# The Challenges of Surveying Heavy-tail Distributions for Use in Infectious Disease Dynamics

**DOI:** 10.1101/2023.07.05.23292248

**Authors:** Michael E. DeWitt, Nicholas Kortessis, John W. Sanders, Candice J. McNeil

## Abstract

Sexual networks often have heavy-tails, where a small number of exceptional individuals in a population have many more sexual partners than the average (e.g., more than five standard deviations). Heavy-tails pose challenges when surveying this group, as these exceptional individuals are uncommon in the population (and so hard to detect), but have disproportionate impact on epidemiological questions, such as those related to the spread of sexually transmitted diseases. In essence, omitting these individuals is a severe error. In this modeling study, we use prior estimates of the distribution of sexual partners amongst men who have sex with men to explore the implication of different sample sizes on survey estimates. We find that even large surveys consistently fail to capture the variance of the sexual network. Surveys of heavy-tailed sexual networks should be designed with this high variance in mind so as not to underestimate the disease dynamics. The failure to adequately capture the variance within a heavy-tailed network has strong implications for infectious disease dynamics and modeling as disease dynamics are often driven by the heavy-tail.

## Introduction

In early 2022, mpox (formerly known as monkeypox) emerged out of traditionally endemic areas of eastern and central Africa and was identified across the world [1]. Monkeypox virus, the virus which causes mpox, was traditionally thought to have lower epidemic potential due to low rates of human-to-human transmission [2]. Human-to-human transmission typically occurs by close and prolonged skin-to-skin contact, in addition to fomites and respiratory droplets [3]. Prior epidemiological analysis and modeling studies had estimated basic reproduction numbers less than 1, indicating low potential for epidemic growth [2, 4]. However, many gay, bisexual, and other men who have sex with men were being diagnosed after reported sexual contact [5], suggesting sustained mpox spread. Mathematical modeling studies were conducted to explain mpox spread despite the thought that the disease should stutter to a stop. These studies indicated that the highly diffuse sexual networks among men who have sex with men may allow for the virus to achieve a basic reproduction number greater than one [6]. Furthermore, Endo and colleagues have referred to their findings as evidence of “heavy-tailed sexual contact networks”, a concept related to probability distributions in which probability density for larger values is higher than that of the exponential distribution. This phenomena relates to the superspreader phenomena which has been observed in other infectious disease processes where a few infections generate a large number of infections due to large variations in individual infectiousness[7, 8]. In the context of sexual networks, there is extremely high variability between reported sexual partners with a very small proportion of individuals having accounting for most sexual partners (Fig. 1). These few, exceptional individuals, greatly increase the population-level basic reproductive number. The outbreak of mpox has shown that understanding the role of these sexual networks is vital in modeling the potential for infectious disease transmission.

**Figure 1:**
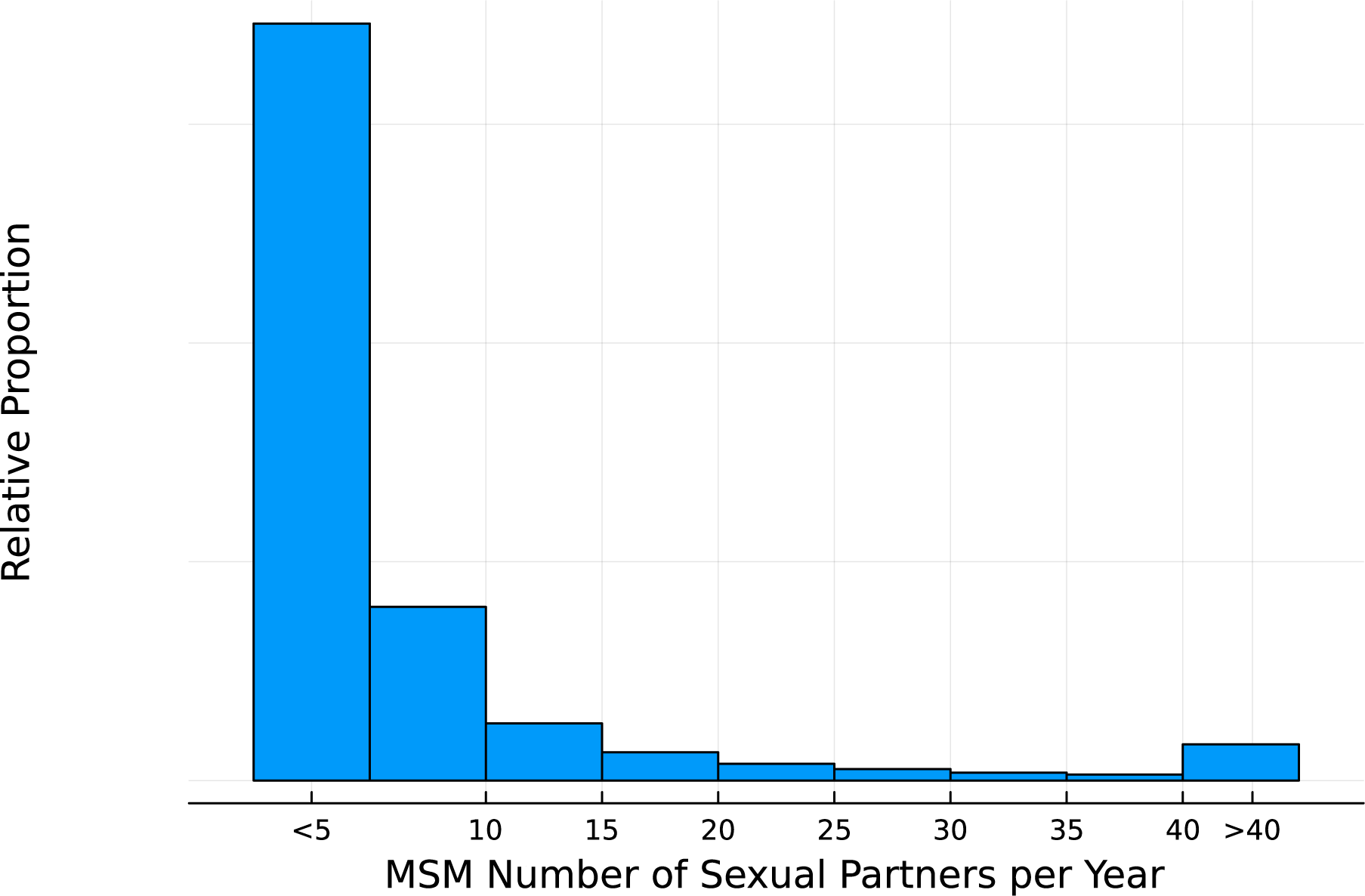
Distribution of sexual partners drawn from a truncated Weibull distribution.

The role of sexual networks on the transmission dynamics of infections and sexually transmitted transmitted infections more specifically has been well understood [9–12]. As more contacts are made, there are more opportunities for infected hosts to transmit the pathogen to their contacts. These insights have been integrated into many mathematical models estimating how infections move within sexual networks [13, 14]. However, many of the data used to generate the network models are limited in the number of participants due to the private nature of the data and the vast heterogeneity in sexual practices. As shown by Endo et al., the heavy-tail of the number of partners complicates understanding these unseen networks [6]. Furthermore, the role that sample size plays in appropriately capturing the heterogeneity of these heavy-tailed networks remains largely unexplored. Achieving proper estimates for modeling sexual networks is vital in developing strategies for disease control and outbreak prevention. In this paper we explore the implications of different sample sizes given existing literature on sexual networks.

## Material and Methods

### Data sources

Following the work of Endo et al. we examine the truncated discrete Weibull distributions which were fitted to empirical data from sexual partnerships from the National Surveys of Sexual Attitudes and Lifestyles (NATSAL) from three survey waves: Natsal-2 (1999–2000; 12,110 participants) (26), Natsal-3 (2010–2012; 15,162 participants) [6, 15]. The NATSAL data includes information on the reported number of sexual partners over a given year for those aged 18-44. In this analysis we considered only the distribution of partners amongst men who have sex with other men (MSM) which were estimated from 409 participants in the combined data sets. The Weibull distribution, a continuous probability distribution with support for positive real numbers, has two parameters: a scale parameter *α* and shape parameter *β*. The shape of the distribution is primarily governed by the scale parameter with values less than one resulting in more mass towards zero while scale values greater than one have more mass in the tails. However, the mass in the tails is greater than that of the exponential distribution. When the scale parameter is one, the Weibull distribution reduces to the exponential distribution, a distribution which is commonly used to model the number of contacts in social networks. In order to account for the discrete nature of sexual partners, we passed the continuous estimates generated from the Weibull distribution through a Poisson distribution in order to account for the discrete nature of sexual contacts. We utilized the parameter estimates for MSM sexual networks from Endo et al. in (Table 1). These parameters result in a mean number of annual sexual partners of 9.24 amongst MSM, however, nearly 70% of the population has fewer than 5 partners, but nearly 5% have more than 30 (Fig. 1). The estimates for this distribution are broadly consistent with the findings from the 4,904 participant study by Weiss and colleagues who found a mean number of annual active partners of 8.5 [13].

**Table 1:** Weibull distribution parameters estimates from Endo et al.

|  |  |
| --- | --- |
| $\alpha$ | $\beta$ |
| 0.1 | 1.36e-9 |

### Estimation of survey representativeness

To understand the role of sample size on estimates of the number of sexual partners within an MSM network, we generated a synthetic population of 100,000 participants using a truncated Weibull distribution to represent the population of MSM and their number of sexual partners in a given geography. Similar methods have been described previously [6, 16] to approximate the distribution of contacts. We then sampled this synthetic population with increasing sample sizes and calculated summary statistics. This process was repeated for 1,000 different synthetic populations.

For each sample size, we calculated several summary statistics including the statistical coverage of the true mean number of sexual partners, the mean absolute error (MAE), and mean absolute percent error (MAPE). In order to assess the statistical coverage, the sampled values were fit using a Poisson model and the estimated rate parameter was recorded. The use of the Poisson distribution for contacts approximates an Erdos-Renyi model for the distribution of contacts in a random network [17]. Erdos-Renyi models are often used to parameterize network models for random network connections an have been used extensively in modeling sexual partnerships [13, 18]. To examine the results in the context of epidemiological estimates, we then used the properties of the sample to estimate the basic reproduction number, *R*_0_, given a recover rate, *γ*, of 10 days and a contact rate, *β*, of 0.1%. Assuming that sexual partnerships are formed at random and in proportion to their count, we utilized the methodology described in Keeling and Rohani to estimate the contact rate using the mean, **M**, and variance, **V**, of the number of contacts in the network as shown in equation 1 [19].

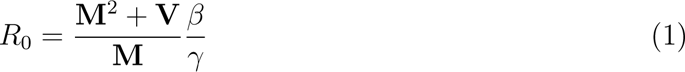

When generating synthetic populations, the heavy-tailed distribution was observed in the quantity of individuals who have a greater number of partners(Figure 1).

## Results

We found that poor coverage of the true mean number of partners was observed under all sampling strategies (Figure 2).

**Figure 2:**
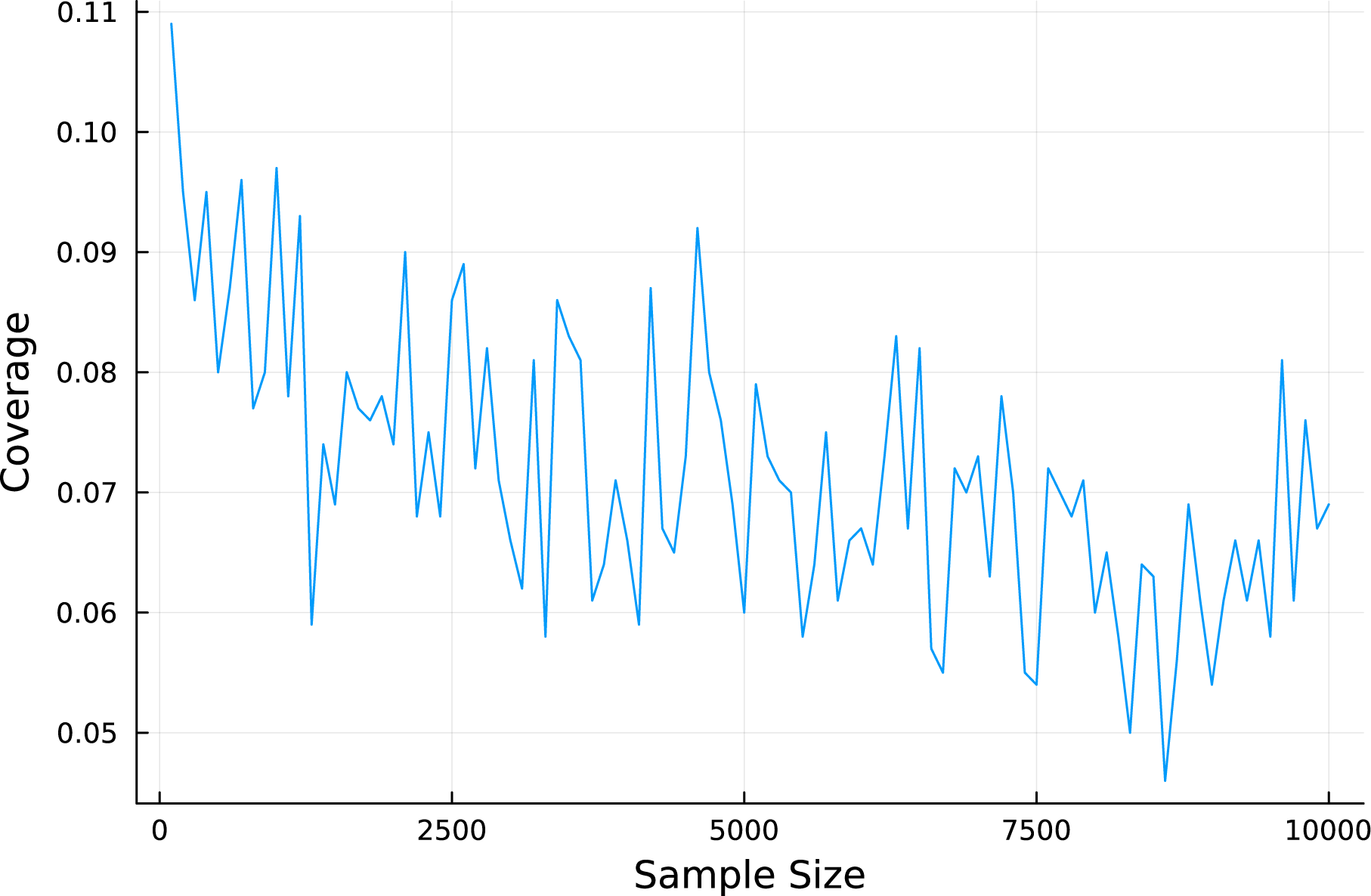
Statistical coverage of the true mean number of sexual partners in the synthetic population based on survey sample size.

When examining the estimates, we find that surveys with smaller sample sizes consistently underestimated the true number of sexual partners (Figure 5, panel A). Similarly, because of this underestimation of the mean number of partners as associated smaller estimated variance, the true basic reproduction number was generally underestimated (Figure 5, panel B). In essence, it is much more likely to sample individuals with below average number of sexual contacts than above average sexual contacts, which consistently biases estimates of the mean to be lower than the true value.

When investigating the average expected bias of the estimate across all iterations of a given sample size, we found that there was no clear convergence to a single value for bias (Figure 4, panel A). Similarly when examining the average proportion of the true variance captured compared to the true variance, we find that the variance was consistently underestimated (Figure 4, panel B).

**Figure 3:**
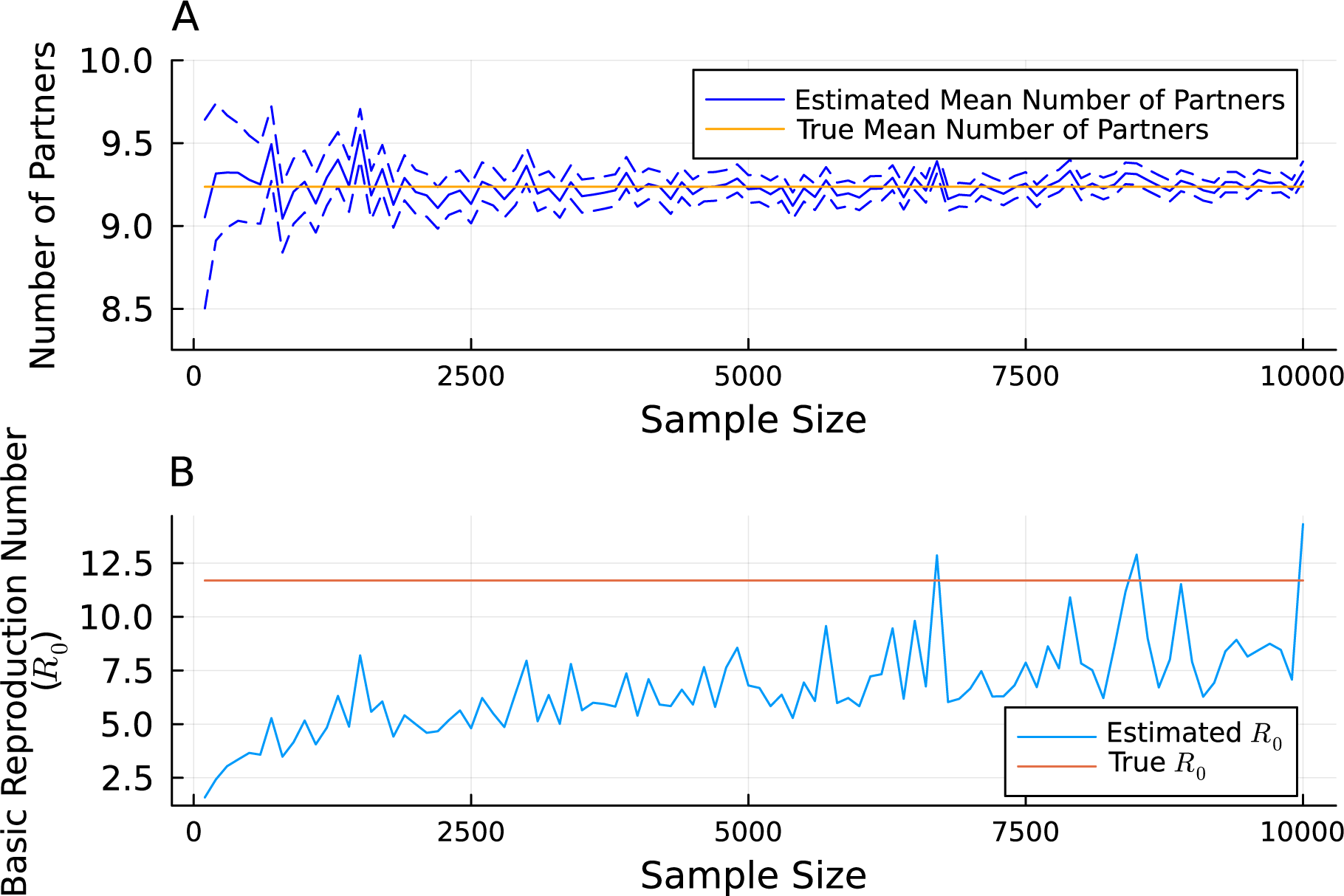
The estimated and true values of the mean number of sexual partners, 95% confidence intervals from a Poisson regression shown (A). The calculated and true contact rate given synthetic epidemiological parameters (B).

**Figure 4:**
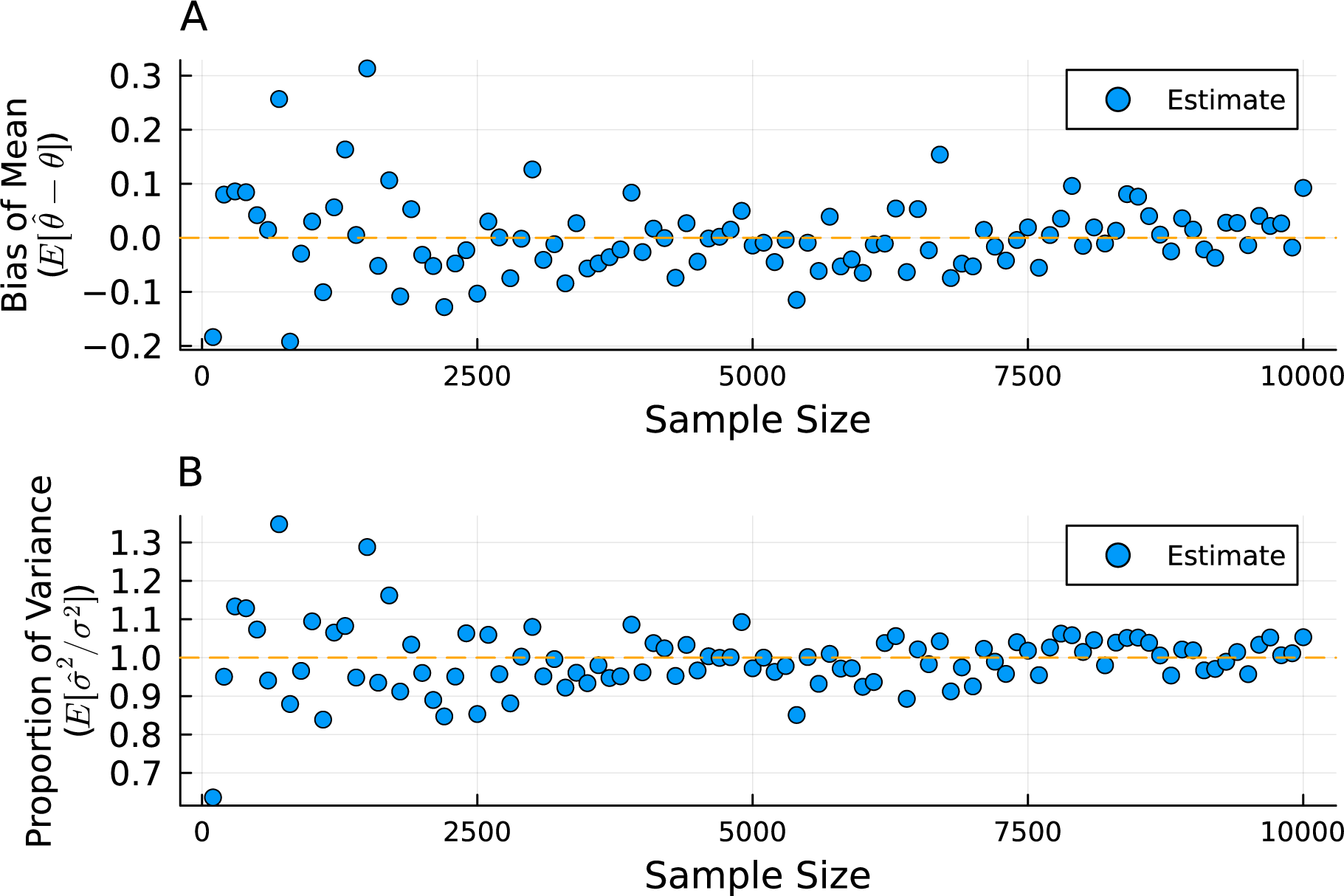
The mean value of the bias for the average number of sexual partners based on the sample size (A). The mean proportion of the variance in the number of sexual partners recovered as a function of the sample size (B).

However as the sample size increased both the mean absolute error and the mean absolute percent error began to converge after samples of 2,000 individuals were reached (Figure 3, panels A and B).

## Discussion

In our study of different surveying approaches of heavy-tailed networks, we found poor statistical coverage of the mean number of partners. We found that the average error, both on absolute (MAE) and proportional scales (MAPE) decreased rapidly after 2,000 individuals were sampled. Given the poor statistical coverage and consistent underestimation of the true variance of the network, estimates of the basic reproduction number for contact networks expected to have heavy tails are bound to be biased far below their true value. And the heavier the tail, the larger the bias in estimation of the basic reproductive number specifically and transmission rates more generally. In essence, the individuals most influential on transmission are scarce in the population and are unlikely to be sampled.

These findings suggest that when surveying heavy-tailed social networks, many more individuals need to be sampled than is standard in most approaches assuming distributions closer to normal. As such, care must be taken to sample a sufficient number of the population in order to properly characterize these networks. These findings have strong implications for future field surveillance and modeling efforts.

### Challenges of heavy-tailed distributions

A key finding of this analysis is that because of extreme events, here sexual partners, the mean degree of the network will be poorly measured if insufficient samples of the population are taken. As demonstrated in our modeling study, this is further complicated by the fact that there did not appear to be convergence to a consistent value of bias as the sample size increase. Additionally, because of the large samples required to capture comparatively rare, extreme events, the variance estimates in smaller samples will fail to capture the true, larger variance of the population. As the proportion of the population in the heavy-tail of the distribution increases, the total variance will increase. Thus for the same estimator, more samples will be required in heavy-tailed distributions than distributions with thinner tails. Use of the Poisson distribution to model these effects will result in overconfidence in the uncertainty surrounding the estimates of the rate parameter due to the high variance of the true parameter despite growing sample sizes.

### Infectious disease dynamics

Early studies of the dynamics of sexually transmitted infections in social networks resulted in discovery of reservoirs of infectious diseases due to heterogeneous mixing patterns of partners [20, 21]. Dense, heavy-tailed sexual networks allow for higher persistence of infection due to their scale free nature [22–24]. This is in contrast to so called “small-world” networks which are best displayed as ring lattice where local connections tends to die out because of local pockets of immunity [16]. Additionally, sexual networks have been shown to be highly diffuse with partners traveling across domestic and international boundaries [25, 26]. Providing reliable estimates for these networks is vital in forming interventions to reduce the likelihood of outbreaks and controlling infection. Importantly, the role these networks play extends to understanding the dynamics of multi-drug resistance organisms such as *Neisseria gonorrhoeae* [27] and sexually transmitted *Shigella flexneri* [28]. It is because of the long tails of these distributions that there is a higher potential risk for explosive outbreaks and persistence with the human population.

Better characterization of these networks could be performed to supplement the information regarding the number of partners. Information about demographics, travel history, and other potential features of a given population could be used to supplement partner information to better capture the true network dynamics. For example, knowledge of participation in group sexual activities could allow modelers to better capture information about the long tail of sexual activities, estimate this population size and thus inform not only survey approaches but also better understand how disease may move within the network.

### Implications for modelers

These findings suggest that higher rates of survey participants may be needed in order to more fully capture the dynamics of these heavy-tailed network. Because the infectious disease dynamics of these networks is largely driven by the heavy-tail of the distribution is it vital that care is taken to adequately capture the bias and variance. As we have shown in our simulation, depending on the sample size, the estimated basic reproduction number from a sample for a pathogen could be 5 times lower than the true value (Figure 5, panel A). This has large implications for the anticipated number of cases, potential number of hospitals, and depending on the virulence, the number of deaths. Furthermore, different control strategies might be adopted with information regarding the basic reproduction number [29].

**Figure 5:**
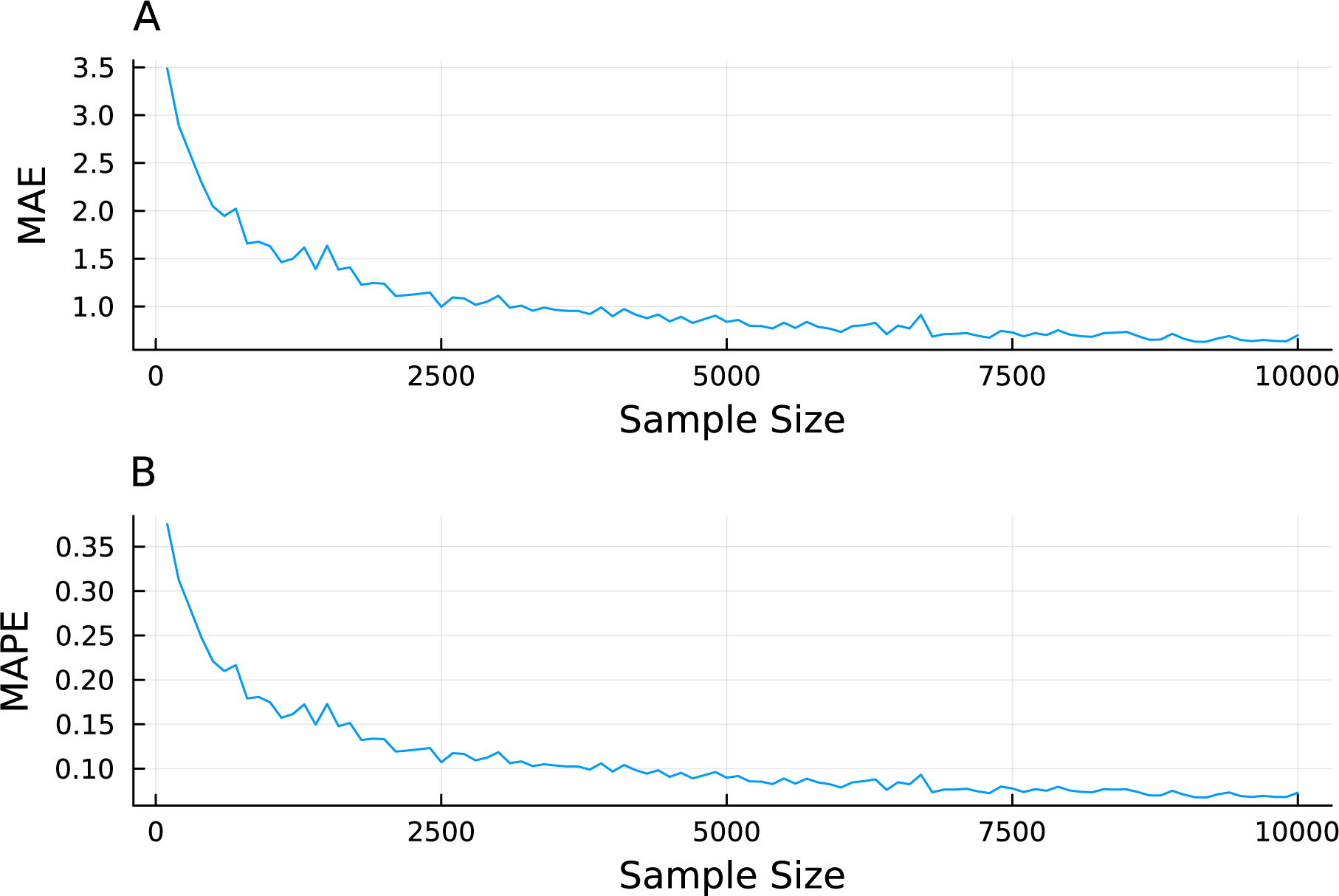
The estimated and true values of the mean number of sexual partners, 95% confidence intervals from a Poisson regression shown (A). The calculated and true contact rate given synthetic epidemiological parameters (B).

Furthermore, modeling studies have shown that when risk-based models are used, the binning strategy for risk can have an impact on the inferences [30]. Furthermore, there are likely additional spatial, behavioral, and psycho-social phenomena that may need to be captured in order to capture how these social networks are connected. We did not assess the role of assortative mixing or homophily where those who are more like one another associate more strongly. Studies have shown that outbreak dynamics and effectiveness of different intervention strategies may differ depending on the overall contact structure and the degree of assortative mixing [31, 32]. As more data become available, these demographic, social, and spatial features could be user to better capture the tail behavior in models considering dense sexual networks. In the absence of data, modeler should be sensitive to the fact that existing data may categorically underestimate the true variance in networks and conduct sensitivity analyse to capture the fuller range of possibilities.

## Data availability

All data and code are stored in a publicly available GitHub repository (https://github. com/wf-id/heavy-tail-sampling).

## Author contribution

All authors contributed equally to the research design, data collection, formal analysis, interpretation of the results, and writing of the paper.

## Data Availability

All data and code are stored in a publicly available GitHub repository https://github.com/wf-id/heavy-tail-sampling.

